# A 515,579-Genome Reference Panel Improves Rare-Variant Imputation Across Multiple Underrepresented Populations

**DOI:** 10.64898/2026.08.25.26361247

**Authors:** Franjo Ivankovic, Arthur Ko, M. Morgan Aster, Mary K Balaconis, Eric Banks, Matthew Bemis, Kristian R Cibulskis, Kylee Degatano, Laura D Gauthier, George Grant, Aaron Hatcher, Christopher Kachulis, Konrad J Karczewski, Sofia M Labrecque, Jonathan Lawson, Calwing Liao, Ricky Magner, Ruchi Munshi, Michael C Schatz, Patrick M Schultz, Saloni P Shah, Elizabeth A Sheets, Kathleen Tibbetts, Kyle A Vernest, Robert Ye, Stacey Gabriel, Niall J Lennon, Benjamin M Neale, Brian L Browning, Lee T Lichtenstein

**Author notes:** Corresponding authors: Arthur Ko and Lee T Lichtenstein. These authors contributed equally: Franjo Ivankovic and Arthur Ko. Senior authors: Brian L Browning and Lee T Lichtenstein.

## Abstract

Genotype imputation remains essential for large-scale human genetics studies, but its performance is limited by the size and ancestral diversity of available reference panels, reducing accuracy for rare variants and underrepresented populations. Here, we present a cloud-based imputation service built on a multi-ancestry reference panel derived from 515,579 jointly phased genomes from the *All of Us* (N=414,830) and National Human Genome Research Institute’s *Analysis, Visualization, and Informatics Lab-space* (AnVIL, N=100,749) datasets. The *All of Us +* AnVIL reference panel is highly diverse and includes 261,163 participants most genetically similar to non-European reference populations, spanning 665,398,839 high-quality autosomal sites, representing a nearly 50% increase over TOPMed, the previous largest imputation service. Across multiple ancestry groups, the panel enables accurate imputation (empirical R² > 0.8) for variants with allele frequencies as low as 0.2%, extending reliable imputation into the rare-variant frequency spectrum, including allele frequencies down to 0.002% and 0.006% for samples with European ancestry and African ancestry in the United States, respectively. Compared with TOPMed, the panel improves imputation accuracy across all ancestry groups except Africans, and recovers additional trait-associated variants not represented in existing reference panels. To facilitate broad community access while preserving participant privacy, we deploy the panel through a secure cloud-based imputation platform using privacy-preserving recombined haplotypes. This resource establishes a new foundation for genome-wide association studies (GWAS) and fine-mapping, especially in previously underrepresented populations.

## Main Text

Genotyping arrays are fast and cost-effective tools for profiling the genomes of hundreds and thousands of individuals.^1,2^ Even as the cost of whole-genome sequencing (WGS) continues to drop, arrays are still widely used as they offer the advantage of enabling larger sample sizes and, consequently, stronger statistical power to map trait-associated genetic loci. More than 23 million individuals have used genotyping arrays through direct-to-consumer programs, and millions more have been profiled across large biobanks and precision medicine initiatives, including the Million Veteran Program, UK Biobank, FinnGen, BioBank Japan, and many others.^3–7^ However, many genetic variants, including low-frequency and rare variants, are not genotyped by arrays, and imputation is typically performed to fill in these missing genotypes.

Genotype imputation infers missing or unobserved variants based on linkage disequilibrium (LD) information from a reference panel consisting of a large number of phased genomes.^8,9^ In many analyses, especially GWAS, genotype imputation is now a common preprocessing step, as it increases marker density, enables joint analysis across different genotyping arrays, and improves the power and resolution of association testing and fine-mapping.^9^

While technical advancements in software and imputation methods have improved the efficiency and accuracy of inferred genotypes, imputation accuracy is still hampered by the low ancestral diversity of available reference panels.^10,11^ The accuracy of imputation increases with the size and diversity of the reference panel.^12^ The largest multi-ancestry reference panel for imputation to date is the Trans-Omics for Precision Medicine (TOPMed) panel, version *R3*, representing 445,600,184 autosomal and X-chromosome sites across 133,597 participants.^13,14^

To enhance imputation accuracy for all populations, we present a newly assembled, multi-ancestry reference panel built by joint phasing of the combined data from the *All of Us* Curated Data Repository (CDR) version 8, including 414,830 short-read genome datasets^15^, and an additional 100,749 genomes from the Center for Common Disease Genomics (CCDG) datasets from NHGRI’s Analysis, Visualization, and Informatics Lab-space (AnVIL).^16^ In accordance with the *All of Us* and AnVIL data access policies and permission from the *All of Us* Data and Research Center Security team, we processed and harmonized both *All of Us* and AnVIL data in the cloud platform, Terra.^17^ Overall, the combined panel comprises more than one million haplotypes and is the largest imputation reference panel produced to date.

We examined the genetic diversity based on genetic similarity to reference populations from the Human Genome Diversity Panel (HGDP) and the 1000 Genomes Project (1KGP) (additional details provided in the Supplementary Information). The imputation panel includes 254,416 participants most similar to the European reference population (49%, EUR), 101,982 participants most similar to the African reference population (20%, AFR), 90,553 participants most similar to the Americas (previously referred to as “Admixed American”) reference population (18%, AMR), 13,226 participants most similar to the East Asian reference population (3%, EAS), 9,710 participants most similar to the South Asian reference population (2%, SAS), and 1,065 participants most similar to the Middle Eastern reference population (0.2%, MID), as well as an additional 44,627 participants not in clear proximity to a single reference population (9%, Remaining, REM) (Fig. 1A, Supplementary Table 1).^18^ The 261,163 participants not in the EUR ancestry group in the *All of Us* + AnVIL panel would alone constitute the second largest publicly usable imputation reference panel (Fig. 1B). Altogether, the reference panel contains 665,398,839 high-quality autosomal sites, including 877,866,186 single-nucleotide variants (SNVs) and 112,002,582 insertions/deletions (indels).

**Figure 1.**
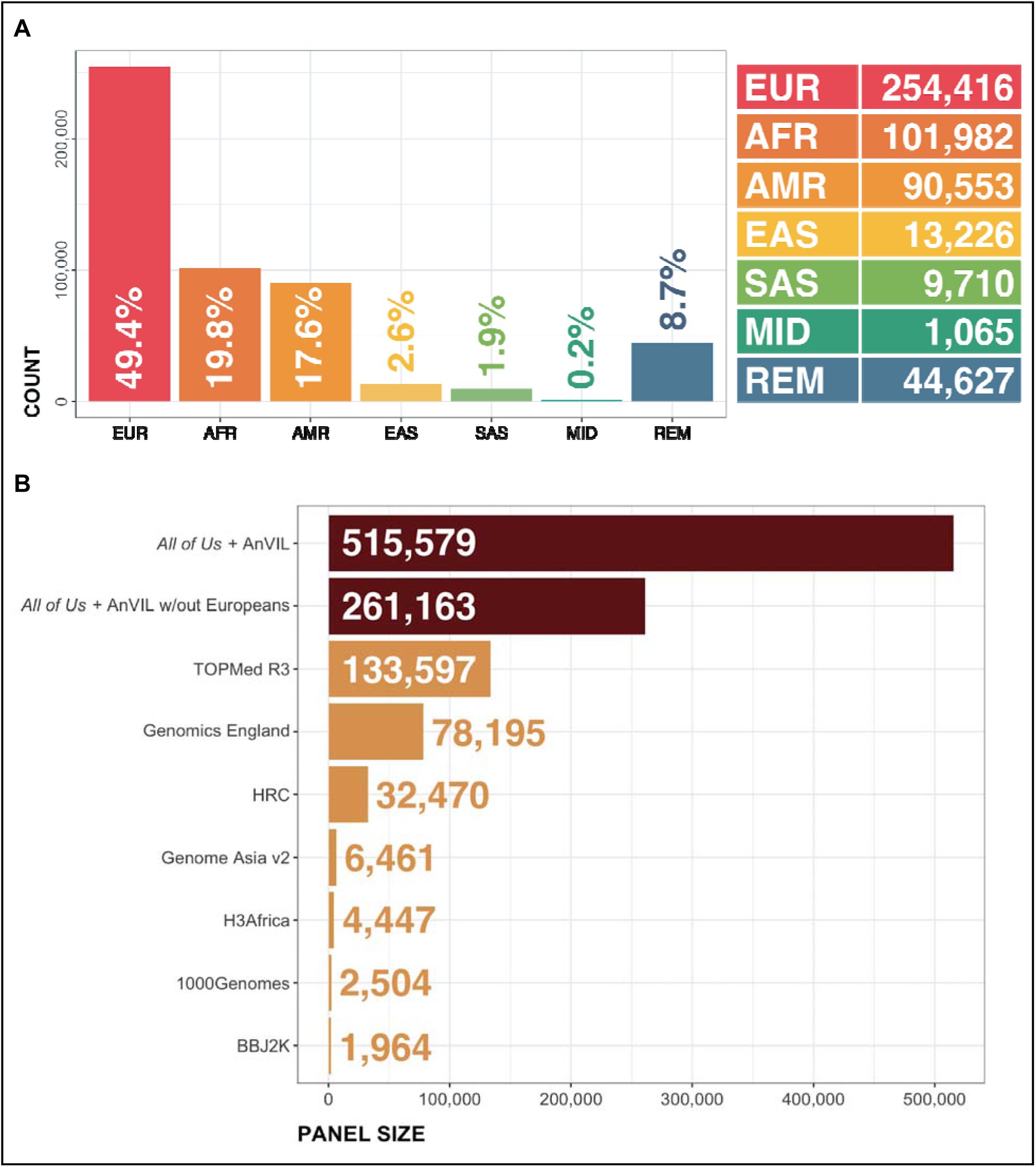
*All of Us* + AnVIL Panel Summary. **A**: Genetic ancestry breakdown of the *All of Us* + AnVIL reference panel, shown as a column plot of participant counts within each major ancestry group: European (EUR; N = 254,416), African (AFR; N = 101,982), Americas (AMR; N = 90,553), East Asian (EAS; N = 13,226), South Asian (SAS; N = 9,710), Middle Eastern/North African (MID; N = 1,065), and Remaining participants (REM; N = 44,627). **B**: Comparison of the complete *All of Us* + AnVIL reference panel size to other commonly used reference panels. The darker bars represent *All of Us* + AnVIL panels with (N = 515,579) and without (N = 261,163) participants with European ancestry. The lighter bars represent other imputation panels, including TOPMed R3^14^ (N = 133,597), Genomics England^28^ (N = 78,195), HRC^29^ (N = 32,470), Genome Asia v2^30^ (N = 6,461), H3Africa^31^ (N = 4,447), 1000 Genomes^32^ (N = 2,504), and BBJ2K^33^ (N = 1,964)

To assemble the panel, we used Hail to perform quality control, filtering, and merging of the *All of Us* and AnVIL data (Supplementary Methods).^19^ Briefly, we first filtered *All of Us* data to exclude sites with > 31 alternate alleles, site-level average sum of allelic depths (AD) < 12, sites with alternate allele count < 2 (singletons and sites without alternate alleles), average site-level call rate < 0.9, and average site-level genotype quality (GQ) mean < 30. We additionally removed sites in *All of Us* annotated with a filter field as having a low quality score (“LowQual”), no high-quality genotypes (“NO_HQ_GENOTYPES”), or excessive heterozygosity (“ExcessHet”).

We then performed a similar quality control in the AnVIL data separately, with a few notable differences. First, we did not filter sites by the number of alternate alleles or alternate allele counts, as this collection is smaller than *All of Us*, so that singletons in the AnVIL data may be more frequent in the combined panel. Second, we selected similar, but not identical, quality-control fields to filter AnVIL data, since *All of Us* used DRAGEN and AnVIL used GATK for variant calling.^20,21^ Finally, we also removed variants from AnVIL datasets not present in the larger *All of Us* dataset. The two individually quality-controlled datasets were then combined using the variant dataset (VDS) combiner function in Hail and additionally filtered to exclude sites with > 31 alternate alleles, prior to exporting to a variant call format (VCF) file for phasing. Phasing was performed on the joint VCF using Beagle v5.5.^8^ The Beagle window-markers parameter was lowered from its default value for most chromosomes to avoid potential out-of-memory errors (Fig. 2). All other Beagle parameters were set to default values.

**Figure 2.**
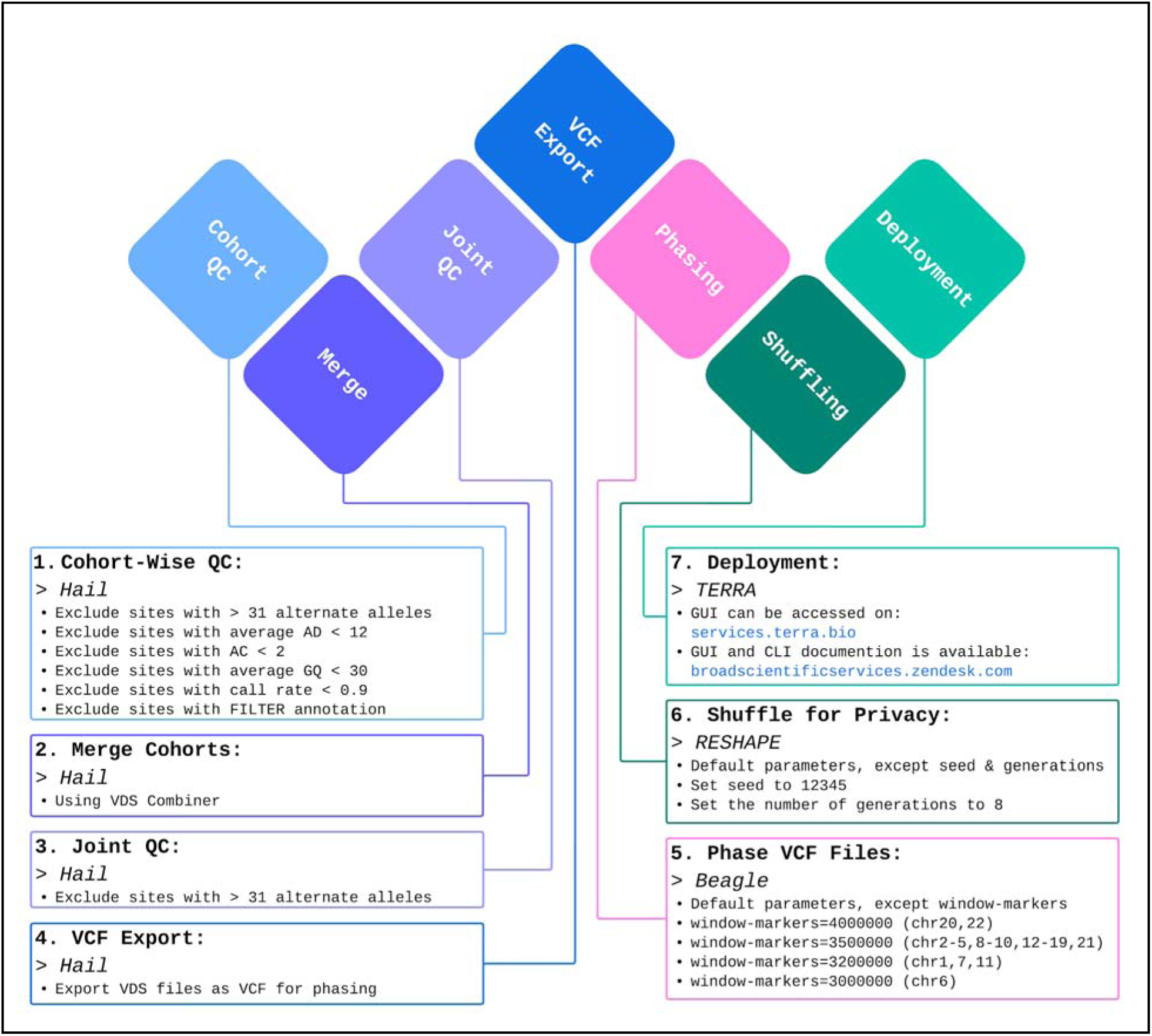
Diagrammatic representation of the *All of Us* + AnVIL reference panel generation and deployment. The diagram depicts steps in the *All of Us* + AnVIL reference panel generation and deployment, including a description of software and parameters used for quality control, phasing, shuffling, and deployment.

To protect the data privacy of *All of Us* and AnVIL participants, we de-identified the phased haplotypes using RESHAPE, which creates recombined genomes by simulating genetic recombination over multiple generations.^22^ RESHAPE was applied to the combined VCF files to simulate 8 generations of recombination events, which was previously shown to have a high probability that each haplotype would recombine at least once per chromosome. Finally, we split 665,398,839 multi-allelic sites into 989,868,768 individual variants, resulting in 877,866,186 SNVs and 112,002,582 indels in the panel.

To evaluate the performance of the *All of Us* + AnVIL reference panel, we benchmarked the imputation for 42 samples with matched genotyping array data (Illumina Global Diversity Array, ∼1.9 million markers) and 30X WGS, which were not included in the imputation panel. This validation set consisted of 32 cell lines from Coriell Institute’s Human Variation Panel (CIHVP) and 10 blood samples purchased from a commercial company, Precision for Medicine. The samples were grouped based on their genetic similarity to 1KGP reference populations, and imputation accuracy was examined for each genetic ancestry group individually, as described in the Supplementary Methods. Initial results showed two subgroups of validation samples most similar to the African reference population, with diverging imputation accuracy. One of these subgroups was composed of cell lines from the Coriell Institute’s *Human Variation Panel - Africans South of the Sahara*. The other subgroup consisted of cell lines from the Coriell Institute’s *Human Variation Panel - African American,* and two blood samples collected in the United States. We therefore reported accuracy for each of these groups separately and referred to them as AFR-SoS and AFR-US, respectively. The number of validation samples in each genetic ancestry group is provided in Supplementary Table 2. We calculated the empirical squared Pearson correlation (ER²) between imputed alternative allele dosages and true alternative allele dosages from WGS data. Filtering by the commonly applied threshold of inferred squared correlation (IR^2,^ which is reported in the “DR^2^” field output by Beagle) > 0.3, 19,114,996 variants were retained for downstream analysis (Table 1). At this IR^2^ threshold, ER^2^ were greater than 0.8 for known SNVs in gnomAD.v4 with allele frequencies as low as 0.2% across all genetic ancestry groups except the East Asian genetic ancestry group (Fig. 3A, Supplementary Table 3). For indels, ER^2^ were greater than 0.8 for variants with allele frequencies > 2% across all genetic ancestry groups except the East Asian genetic ancestry group (Fig. 3B, Supplementary Table 3). In summary, the *All of Us* + AnVIL panel can provide reliable imputation for different ancestry groups, even for low-frequency and rare variants.

**Figure 3.**
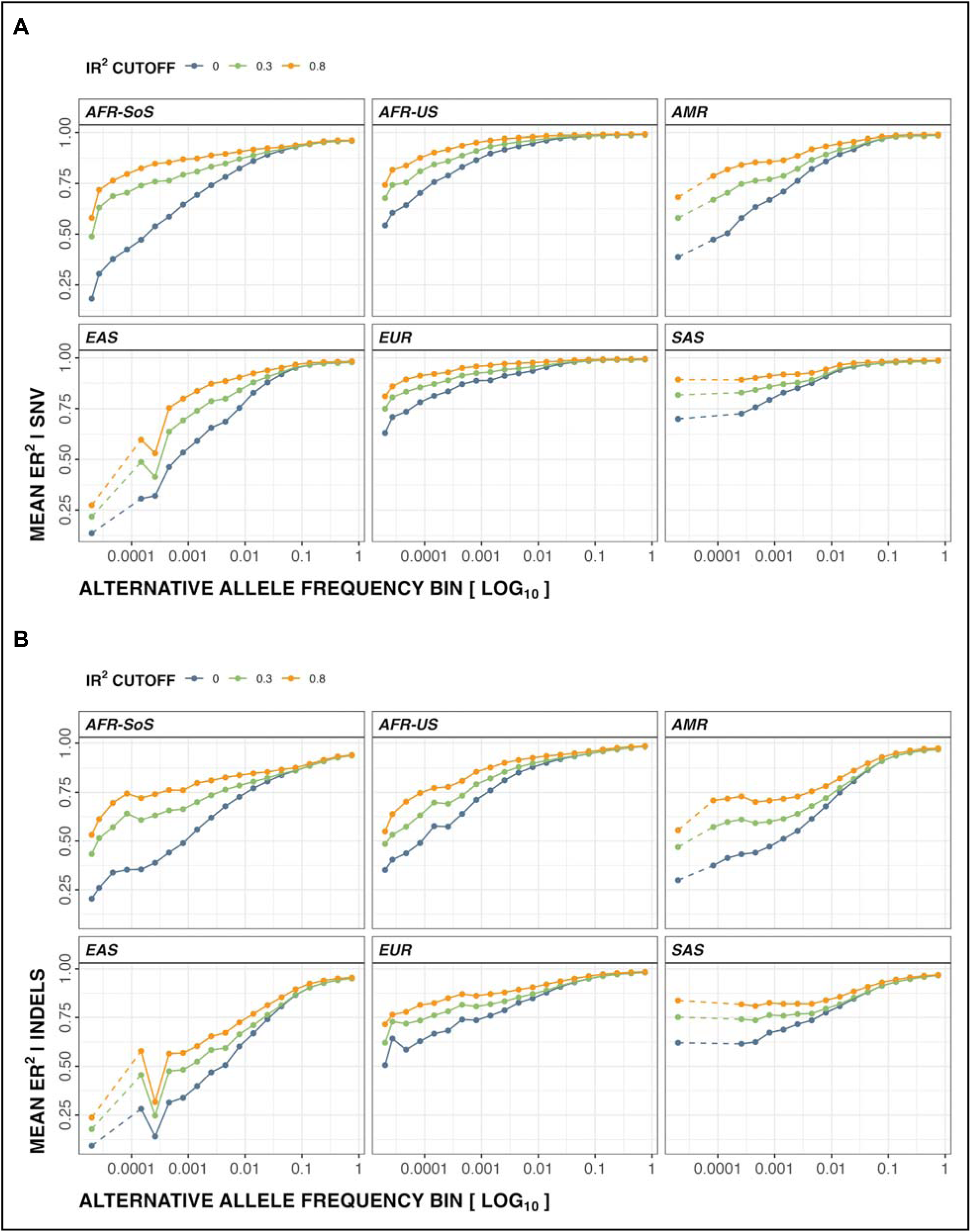
Performance evaluation of the *All of Us* + AnVIL imputation panel. The empirical squared correlations (ER^2^) between imputed allele dosage and WGS allele dosage plotted against allele frequency, stratified by different thresholds of inferred squared correlation (IR^2^) computed by Beagle (shown in color, blue for 0, green for 0.3, and orange for 0.8). The ER^2^ of SNVs are shown in panel (**A**), and indels are shown in (**B**). Variants were binned based on ancestry-specific allele frequencies from gnomAD, and missing frequency bins are shown as dashed lines, likely due to limited WGS sample sizes of the respective ancestry groups in gnomAD. Using IR^2^>0.3 as a cutoff, the *All of Us* + AnVIL panel can reliably impute SNVs (ER^2^>0.8) with frequencies in gnomAD as low as 0.2% across all genetic ancestry validation groups except EAS. With the same IR^2^ filter, indels with allele frequencies >2% can be imputed with high confidence (ER^2^>0.8) for all except EAS. Genetic ancestry group abbreviations of the validation set are defined in Supplementary Table 2.

**Table 1.** The number of imputed variants in 42 CIHVP samples for the *All of Us +* AnVIL and TOPMed panels, stratified by inferred squared correlation (IR^2^) cut-offs.

|  | Number of variants |  | Variants present in both panels |
| --- | --- | --- | --- |
| $IR^2$ Cutoff | *All of Us* + AnVIL | TOPMed | |
| 0 | 989,849,055 | 422,893,758 | 340,043,348 |
| 0.3 | 19,114,996 | 15,998,556 | 14,915,530 |
| 0.8 | 18,133,229 | 14,945,516 | 14,273,202 |

Next, we compared the imputation performance of the *All of Us* + AnVIL reference panel and the TOPMed Reference panel. We used the same validation samples that were not included in either imputation panel and imputed against the *All of Us* + AnVIL reference panel using Beagle. For TOPMed, imputation was performed using the Michigan Imputation Server^23^. We first examined the intersecting variants imputed by both panels, representing around 34.3% of total variants in the *All of Us* + AnVIL reference. (Table 1). After imputation, *All of Us* + AnVIL outperformed TOPMed in five of the six genetic ancestry groups, with the largest gains observed among low-frequency and rare variants (Fig. 4). Specifically, we find >10% ER² improvement for rare variants (0.1% MAF) across AMR, EAS, EUR, and SAS genetic ancestry validation groups. The *All of Us* + AnVIL panel outperforms TOPMed by a similar margin in the AFR-US group, consistent with the substantial size of the AFR reference panel group and recruitment by *All of Us* from within the United States and its territories. However, TOPMed achieved higher accuracy for the AFR-SoS group, potentially because TOPMed includes more participants who are genetically more similar to those in the AFR-SoS group.^14^ Overall, our comparative analysis highlights the need for diverse and representative panels, and that expanding large, ancestrally diverse reference resources can improve rare-variant imputation across globally distributed populations.

**Figure 4.**
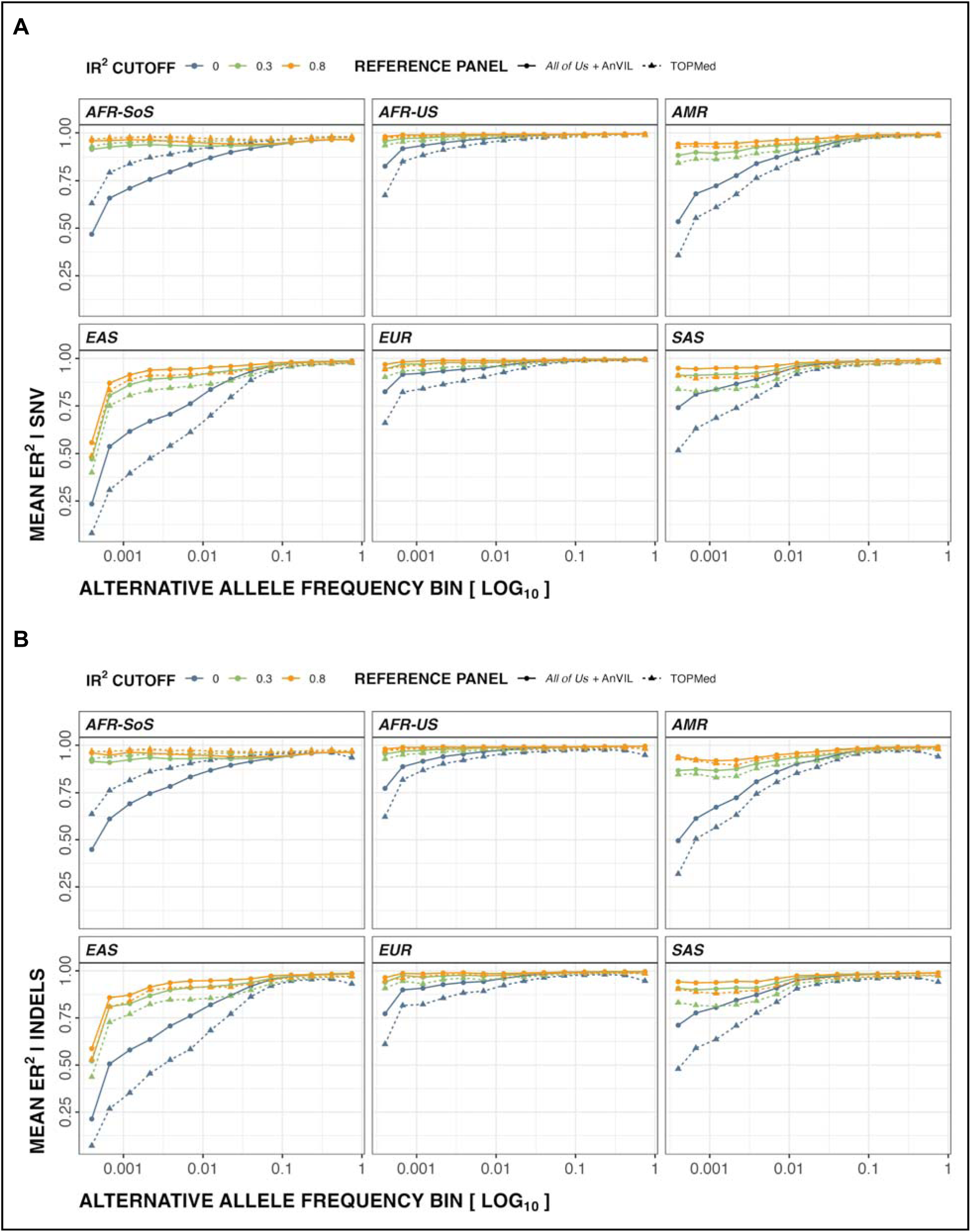
Comparison of TopMed and *All of Us* + AnVIL imputation performance. Comparison of imputation performance for SNVs **(A)** and Indels **(B)**. Colors show IR^2^ stratification (blue for 0, green for 0.3, and orange for 0.8), and linetypes/point shapes show reference panels (circle points on solid lines for *All of Us* + AnVIL and triangle points on dashed lines for TOPMed). *All of Us +* AnVIL provides higher overall accuracy than TOPMed across all ancestry groups except AFR-SoS. Genetic ancestry group abbreviations are defined in Supplementary Table 2.

We then assessed how the new imputation panel could improve GWAS power relative to TOPMed. We analyzed GWAS hits for 34 complex traits and diseases, including height, Type 2 diabetes, and hypertension, from a recent study of European participants found by WGS in the U.K. Biobank^24^. This study identified 243 unique trait-associated variants that were captured by WGS but missed by TOPMed imputation in the original GWAS analyses. We examined the imputation outputs from both *All of Us* + AnVIL and TOPMed and found that 46.5% of the reportedly missed variants were present in both imputation panels and were likely excluded from the original study in the U.K. Biobank due to downstream quality controls. Furthermore, roughly 42% of the variants reside within low-complexity regions (LCRs) that were excluded by TOPMed’s quality control^25,26^. Of the remaining 28 variants, six were not present in either panel, one was exclusive to TOPMed, and one variant was excluded by both panels and was marked as a low-quality site by gnomAD. The remaining 20 were uniquely found by the *All of Us* + AnVIL panel, including 13 common and 7 rare variants not present in TOPMed. Overall, the *All of Us* + AnVIL panel provides 20 additional common and rare genomic variants with known associations from the U.K. Biobank study, compared to TOPMed.

Finally, we introduce the *All of Us* + AnVIL Imputation Service, a new cloud-based service that enables users to quickly and securely perform phasing and imputation against this new reference panel (Fig. 5). The service is hosted on the Terra platform with *All of Us*- and NHGRI- compliant FedRAMP/NIST 800-53 Rev. 5 Moderate risk management^27^, with Google Cloud Platform as the backend. Users can interact with the imputation service via either the Python-based command-line tool, *terralab*, or the graphical user interface on Terra (https://allofus-anvil-imputation.broadinstitute.org) to facilitate data transfer and job management. Importantly, the reference panel is de-identified through simulated recombination, and end-users do not have access to *All of Us* + AnVIL individual-level genotype data. Users’ data uploaded to the service are protected by secure monitoring and authorization domains implemented in Terra, and reside within the security perimeter for no more than 14 days before deletion. We tested the performance of the imputation service with 2,000, 5,000, and 10,000 array input samples, and the total runtimes were 22, 42, and 75 hours, respectively, indicating nearly linear time complexity with respect to sample size, given the same computing environment and resources.

**Figure 5.**
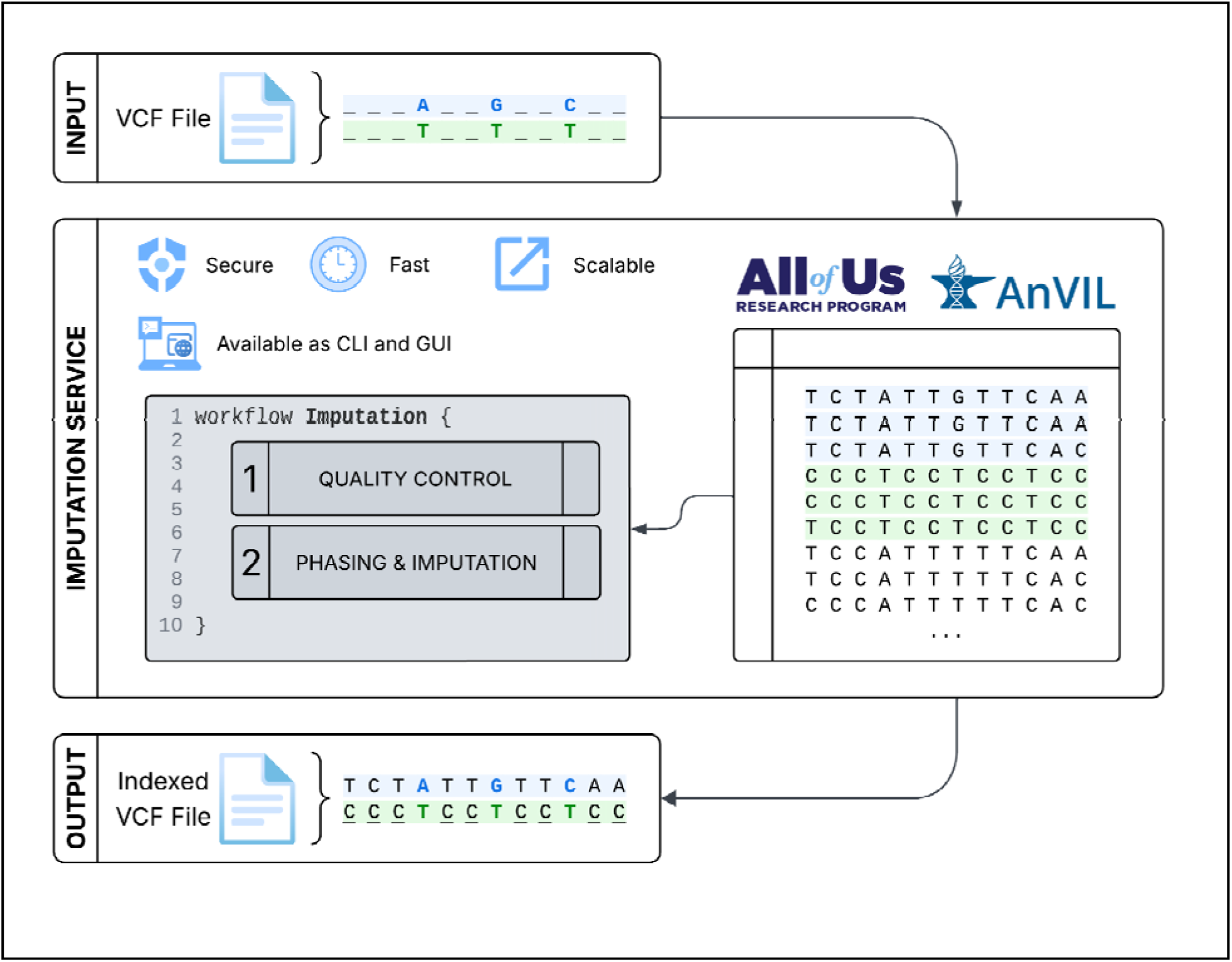
Overview of the imputation service workflow. Users provide a VCF file with genotypes in the secure environment. The imputation workflow automatically performs quality control, phasing, and imputation using the *All of Us* + AnVIL reference panel, and returns an indexed VCF file with imputed genetic data.

The new cloud-based service provides a secure and simple interface for users to access this expansive and diverse panel while protecting participants’ genetic data privacy. One current limitation of the service is its restriction to autosomal chromosomes; we anticipate including the X chromosome in a later update. Furthermore, imputation for low-pass whole-genome sequence data is under development to further enhance the utility of the reference panel. Finally, additional datasets from *All of Us* and AnVIL may be ingested into the panel in the future to further increase the panel’s size and diversity.

## Ethics declarations

K.J.K. is a member of the scientific advisory board of Nurture Genomics. B.M.N is a member of the scientific advisory board at Deep Genomics, Vesalius Therapeutics, Camp4 Therapeutics, and Aluco BioSciences Inc. The remaining authors declare no competing interests.

## Data Availability

All data are or will be available through the All of Us Researcher Workbench or the NHGRI Genomic Data Science Analysis, Visualization, and Informatics Lab-space (AnVIL) under controlled access.

https://allofus-anvil-imputation.broadinstitute.org/

## Acknowledgements

A.K. acknowledges the support from the NIH Office of Data Science Strategy and the National Human Genome Research Institute through the Data and Technology Advancement National Service (DATA) Scholar Program. B.L.B was supported by the National Human Genome Research Institute of the National Institutes of Health, under Award Number R01HG014458. The imputation panel generation and service were supported by awards from the *All of Us* (OT2OD038121, OT2OD002748, OT2OD038111, and OT2OD002750) and the National Human Genome Research Institute (5U24HG010262 and 5U24HG010263). B.M.N. was supported by the National Institute of Mental Health (R37MH107649). The content is solely the responsibility of the authors and does not necessarily represent the official views of the National Institutes of Health. We thank the TOPMed imputation service team and the AnVIL team for helpful discussion and suggestions. The Centers for Common Disease Genomics (CCDG) are funded by the National Human Genome Research Institute (UM1HG008853, UM1HG008895, UM1HG008898, UM1HG008901, and U24HG008956) and in part by the National Heart, Lung, and Blood Institute. We thank the participants and investigators of CCDG for their contributions to the imputation reference panel. Complete and detailed acknowledgments of all CCDG cohorts are listed in the Supplementary Information. We gratefully acknowledge *All of Us* participants for their contributions, without whom this research would not have been possible. We also thank the National Institutes of Health’s *All of Us* Research Program for making available the participant data for this study.

