## Supplementary Information for "A 515,579-Genome Reference Panel Improves Rare-Variant Imputation Across Multiple Underrepresented Populations"

### Supplementary Methods

**Genotype and variant quality control:**
To determine the threshold for each genotype and variant QC metric, we performed phasing accuracy tests in which we excluded trio parents and filtered markers across a range of thresholds for each QC metric prior to statistical phasing. In each test, we recorded the number of excluded markers and the phase accuracy in the trio offspring relative to the phase determined by the parental genotypes. The filtering criteria were chosen to achieve high genotype phasing accuracy while retaining as many markers as possible.

**Genotype concordance analysis**:
We evaluated the performance of imputation by calculating the squared Pearson's correlation ($R^2$) between alternative-allele dosages from imputed variants and WGS calls for 42 Coriell Institute's Human Variation Panel samples. We used the function, "EvaluateGenotypingPerformance," implemented in GATK. Variants were first binned into allele-frequency tranches based on ancestry-specific allele frequencies from gnomAD.v4. Then, variants were aggregated within each bin to calculate $R^2$ for each sample. We used gnomAD as an independent dataset for allele-frequency annotation to avoid bias toward either TOPMed or *All of Us* + AnVIL.

**Genetic ancestry inference**:
Ancestry inference for CCDG participants was performed by the CCDG working group, and the population labels are available on AnVIL. Two pipelines were developed to perform inference using two reference panels: 1) Human Genome Diversity Panel (HGDP) + 1000 Genomes Project (1KGP), and 2) gnomAD v3. The CCDG participants were jointly called with 3,943 HGDP + 1KGP samples, and related participants were initially filtered. Principal component analyses (PCA) were then performed on the joint callset, and related participants were projected back into the PC space. A random forest (RF) classifier was trained using the first 10 PCs and the known population labels from HGDP+1KGP. To confidently predict a participant's genetic ancestry, a 90% probability from the RF classifier was required; otherwise, a participant was assigned to Remaining (REM). The second pipeline. CCDG participants were projected into the PC space of gnomAD.v3 genomes, and an RF model was similarly trained on the PCs and known population labels of gnomAD. A 90% probability threshold was also applied.

Genetic ancestry inference for the *All of Us* participants was previously published (1). Briefly, the HGDP + 1KGP dataset was used to train a random forest classifier using the first 16 PCs derived from high-quality autosomal sites. Then, the *All of Us* participants were projected onto the PCA space of the training data and classified by the random forest model.

Ancestry inference for the validation dataset was performed by projecting samples onto 1KGP principal components, and visually comparing PC1 and PC2 positions of validation samples with 1KGP reference population samples. We further broke down the categories based on the cell line categorization and imputation accuracy. There were two subgroups of validation samples most similar to the 1KGP African reference population but with diverging imputation accuracy. One of these subgroups was composed of cell lines from the Coriell Institute's *Human Variation Panel - Africans South of the Sahara*. The other subgroup consisted of cell lines from the Coriell Institute's *Human Variation Panel - African American*, and two blood samples collected in the United States. We therefore reported accuracy for each of these groups most similar to the African reference population separately, and referred to them as AFR-SoS and AFR-US, respectively.

### Supplementary Tables

**Supplementary Table 1. Genetic ancestry groups and their counts in the *All of Us* and AnVIL imputation panel.**

| **Genetic ancestry group** | **Group acronym** | **Population name** | **Count** |
|---|---|---|---|
| 1KGP-HGDP-AFR-like | AFR | African | 101,982 (19.8%) |
| 1KGP-HGDP-AMR-like | AMR | Americas | 90,553 (17.6%) |
| 1KGP-HGDP-EAS-like | EAS | East Asian | 13,226 (2.6%) |
| 1KGP-HGDP-EUR-like | EUR | European | 254,416 (49.3%) |
| 1KGP-HGDP-MID-like | MID | Middle Eastern | 1,065 (0.2%) |
| 1KGP-HGDP-SAS-like | SAS | South Asian | 9,710 (1.89%) |
| Remaining participants | REM | Remaining | 44,627 (8.7%) |
| Total | - | - | 515,579 |

**Supplementary Table 2. Genetic ancestry group breakdown of the validation samples. Additional method details provided in Supplementary Methods.**

| **Genetic ancestry groups of validation samples** | **Sample size** | **Description** |
|---|---|---|
| AFR-SoS | 4 | Samples from the Coriell Institute's *Human Variation Panel - Africans South of the Sahara* genetically most similar to the 1KGP AFR reference population. |
| AFR-US | 10 | Eight samples from the Coriell Institute's *Human Variation Panel - African American* plus two blood |

| | | |
|---|---|---|
| | | samples collected in the United States, genetically most similar to the 1KGP AFR reference population. |
| AMR | 10 | Nine samples from the Coriell Institute's *Human Variation Panel - South America (Andes)* plus one blood sample collected in the United States, genetically most similar to the 1KGP AMR reference population. |
| EAS | 4 | Three samples from the Coriell Institute's *Human Variation Panel - Chinese,* plus one blood sample collected in the United States, genetically most similar to the 1KGP EAS reference population. |
| EUR | 6 | Blood samples collected in the United States, genetically most similar to the 1KGP EUR reference population. |
| SAS | 8 | Samples from the Coriell Institute's *Human Variation Panel - Southeast Asian (Excluding Japanese and Chinese),* genetically most similar to the 1KGP SAS reference population. |
| **Total:** | **42** | |

**Supplementary Table 3**. **Tabulated empirical $R^2$ ($ER^2$) validation summaries**. The table is attached as an excel spreadsheet titled "Supplementary Table 3".

### Additional Acknowledgements

Variant calls and underlying sequence data of the CCDG cohorts were funded by NHGRI CCDG awards to Washington University in St. Louis (WU) (UM1HG008853), Broad Institute of MIT and Harvard (UM1HG008895), Baylor College of Medicine (UM1HG008898), and New York Genome Center (UM1HG008901); and NHGRI Genome Sequencing Program (GSP) Coordinating Center grant to Rutgers (U24 HG008956).

The following cell lines/DNA samples were obtained from the NIGMS Human Genetic Cell Repository at the Coriell Institute for Medical Research: [NA06984, NA06985, NA06986, NA06989, NA06994, NA07000, NA07037, NA07048, NA07051, NA07056, NA07347, NA07357, NA10847, NA10851, NA11829, NA11830, NA11831, NA11832, NA11840, NA11843, NA11881, NA11892, NA11893, NA11894, NA11918, NA11919, NA11920, NA11930. NA11931, NA11932, NA11933, NA11992, NA11994, NA11995, NA12003, NA12004, NA12005, NA12006, NA12043, NA12044, NA12045, NA12046, NA12058, NA12144, NA12154, NA12155, NA12156, NA12234, NA12249, NA12272, NA12273, NA12275, NA12282, NA12283, NA12286, NA12287, NA12340, NA12341, NA12342, NA12347, NA12348, NA12383, NA12399, NA12400, NA12413,, NA12414, NA12489, NA12546, NA12716, NA12717, NA12718, NA12748, NA12749, NA12750, NA12751, NA12760, NA12761, NA12762, NA12763, NA12775, NA12776, NA12777, NA12778, NA12812, NA12813, NA12814, NA12815, NA12827, NA12828, NA12829, NA12830, NA12842, NA12843, NA12872, NA12873, NA12874, NA12878, NA12889, NA12890]. These data were generated at the New York Genome Center with funds provided by NHGRI Grant 3UM1HG008901-03S1.

The following cell lines/DNA samples were obtained from the NIGMS Human Genetic Cell Repository at the Coriell Institute for Medical Research: [NA06984, NA06985, NA06986, NA06989, NA06991, NA06993, NA06994, NA06995, NA06997, NA07000, NA07014, NA07019, NA07022, NA07029, NA07031, NA07034, NA07037, NA07045, NA07048, NA07051, NA07055, NA07056, NA07340, NA07345, NA07346, NA07347, NA07348, NA07349, NA07357, NA07435, NA10830, NA10831, NA10835, NA10836, NA10837, NA10838, NA10839, NA10840, NA10842, NA10843, NA10845, NA10846, NA10847, NA10850, NA10851, NA10852, NA10853, NA10854, NA10855, NA10856, NA10857, NA10859, NA10860, NA10861, NA10863, NA10864, NA10865, NA11829, NA11830, NA11831, NA11832, NA11839, NA11840, NA11843, NA11881, NA11882, NA11891, NA11892, NA11893, NA11894, NA11917, NA11918, NA11919, NA11920, NA11930, NA11931, NA11932, NA11933, NA11992, NA11993, NA11994, NA11995, NA12003, NA12004, NA12005, NA12006, NA12043, NA12044, NA12045, NA12046, NA12056, NA12057, NA12058, NA12144, NA12145, NA12146, NA12154, NA12155, NA12156, NA12234, NA12239, NA12248, NA12249, NA12264, NA12272, NA12273, NA12274, NA12275, NA12282, NA12283, NA12286, NA12287, NA12329, NA12335, NA12336, NA12340, NA12341, NA12342, NA12343, NA12344, NA12347, NA12348, NA12375, NA12376, NA12383, NA12386, NA12399, NA12400, NA12413, NA12414, NA12485, NA12489, NA12546, NA12707, NA12708, NA12716, NA12717, NA12718, NA12739, NA12740, NA12748, NA12749, NA12750, NA12751, NA12752, NA12753, NA12760, NA12761, NA12762, NA12763, NA12766, NA12767, NA12775, NA12776, NA12777, NA12778, NA12801, NA12802, NA12812, NA12813, NA12814, NA12815, NA12817, NA12818, NA12827, NA12828, NA12829, NA12830, NA12832, NA12842, NA12843, NA12864, NA12865, NA12872,

NA12873, NA12874, NA12875, NA12877, NA12878, NA12889, NA12890, NA12891, NA12892]. These data were generated at the New York Genome Center with funds provided by NHGRI Grants 3UM1HG008901-03S1 and 3UM1HG008901-04S2 (2).

The following DNA samples were obtained from the NHGRI Sample Repository for Human Genetic Research at the Coriell Institute for Medical Research: NA12878, NA12891, NA12892, NA19238, NA19431, NA19648, HG00512, HG00513, HG00514, HG00731, HG00732, HG00733, NA19239, NA19240, HG01350, HG01351, HG01352, HG02059, HG02060, HG02061, HG02816, HG02817, HG02818, NA24143, NA24149, NA24385.

Data was generated as part of the National Institute of Diabetes and Digestive and Kidney Diseases (NIDDK) IBD Genetics Consortium (IBDGC) and International IBD Genetics Consortium (IIBDGC) supported by The Helmsley Charitable Trust and the Centers for Common Disease Genomes Program (NHGRI). DNA samples were obtained from the following collections: The Lunenfeld-Tanenbaum Research Institute, Mount Sinai Hospital, The University of Pittsburgh School of Medicine, The Emory University School of Medicine, The Johns Hopkins Hospital, The Icahn School of Medicine at Mount Sinai, The Washington University School of Medicine, The University of Miami Miller School of Medicine, and Cedars Sinai.

The Hispanic Community Health Study/Study of Latinos was carried out as a collaborative study supported by contracts from the NHLBI to the University of North Carolina (N01-HC65233), University of Miami (N01-HC65234), Albert Einstein College of Medicine (N01-HC65235), University of Illinois at Chicago (HHSN268201300003I), Northwestern University (N01-HC65236), and San Diego State University (N01-HC65237). The following Institutes/Centers/Offices contribute to the HCHS/SOL through a transfer of funds to the NHLBI: National Institute on Minority Health and Health Disparities, National Institute on Deafness and Other Communication Disorders, National Institute of Dental and Craniofacial Research, National Institute of Diabetes and Digestive and Kidney Diseases, National Institute of Neurological Disorders and Stroke, NIH Institution-Office of Dietary Supplements. The views expressed in this manuscript are those of the authors and do not necessarily represent the views of the National Heart, Lung, and Blood Institute; the National Institutes of Health; or the U.S. Department of Health and Human Services.

The CCDG METSIM study was supported by grants from the Academy of Finland (321428), National Institutes of Health, Sigrid Juselius Foundation, and Finnish Foundation for Cardiovascular Research.

We thank contributors who collected samples and/or data used in this study, as well as subjects whose help and participation made this work possible. Biobanking was supported by grants from Aarno Koskelo Foundation, Helsinki University Central Hospital special government funds (EVO #TYH2012209, #TKK2012005, #TYH2014312, #TYH2017250, #TYH2019317), and Finnish Foundation for Cardiovascular Research.

This work was supported by National Institutes of Health (NIH) grants to the Autism Sequencing Consortium (ASC). Sequencing at Broad Institute was supported by the Centers for Common Disease Genomes Program (NHGRI). We acknowledge the clinicians and organizations that contributed to samples used in this study. Finally, we are grateful to the many families whose participation made this study possible.

The NIA-LOAD study supported the collection of samples used in this study through National Institute on Aging (NIA) grants U24AG026395 and R01AG041797. We thank contributors, including the Alzheimer's Disease Centers, who collected samples used in this study, as well as patients and their families, whose help and participation made this work possible.

Genomic data provided by iHART, an initiative led by the Hartwell Foundation.

Data collection for this project was supported by the Genetic Studies of Alzheimer's disease in Caribbean Hispanics (EFIGA) funded by the National Institute on Aging (NIA) and by the National Institutes of Health (NIH) (5R37AG015473, RF1AG015473, R56AG051876). We acknowledge the EFIGA study participants and the EFIGA research and support staff for their contributions to this study.

The TASC consortium collection and genotyping was supported by Autism Speaks. Funding for the sequencing and distribution of TASC samples was provided by the National Institutes of Health (MH094303 and MH100233).

Data collection and sharing for this project were supported by the Washington Heights-Inwood Columbia Aging Project (WHICAP, PO1AG07232, R01AG037212, RF1AG054023) funded by the National Institute on Aging (NIA) and by the National Center for Advancing Translational Sciences, National Institutes of Health, through Grant Number UL1TR001873. We acknowledge the WHICAP study participants and the WHICAP research and support staff for their contributions to this study.

The Atherosclerosis Risk in Communities study has been funded in whole or in part with Federal funds from the National Heart, Lung, and Blood Institute, National Institutes of Health, Department of Health and Human Services (contract numbers HHSN268201700001I, HHSN268201700002I, HHSN268201700003I, HHSN268201700004I, and HHSN268201700005I). The authors thank the staff and participants of the ARIC study for their important contributions.

Samples and associated genotype and phenotype data used in this study were provided by the Center for Applied Genomics at The Children's Hospital of Philadelphia and supported by an Institutional Development Award from The Children's Hospital of Philadelphia and donation from the Lurie Family Foundation. Whole genome sequencing for this project at the New York Genome Center was supported by UM1HG008901. We gratefully thank all the children and their families who enrolled in this study, and all individuals who donated blood samples for research purposes (3).

The collection and analysis of this dataset was supported in part by grants (GNT1044175 and GNT1098255) from the National Health and Medical Research Council (NHMRC) Australia.

We thank all of the families at the participating SAGE study, as well as the principal investigators .

Data was generated as part of the Centers for Common Disease Program (NHGRI) with support from TOPMed (NHLBI). We thank the Broad Institute for high quality sequence data generation and are grateful to our collaborators who have contributed samples for these studies.

This dataset was funded by the Italian Ministry of Health, grant number GR-2013-02357561, and by the University of Bologna, Italy (RFO).

The Mount Sinai BioMe Biobank has been supported by The Andrea and Charles Bronfman Philanthropies and in part by Federal funds from the NHLBI and NHGRI (U01HG00638001; U01HG007417; X01HL134588). We thank all participants in the Mount Sinai Biobank. We also thank all our recruiters who have assisted and continue to assist in data collection and management and are grateful for the computational resources and staff expertise provided by Scientific Computing at the Icahn School of Medicine at Mount Sinai.

Supported by NIH and NHLBI grant # R01HL117004; study enrollment supported by NIEHS grant # R01ES015794, the Sandler Family Foundation, the American Asthma Foundation, the RWJF Amos Medical Faculty Development Program, Harry Wm. and Diana V. Hind Distinguished Professor in Pharmaceutical Sciences II.

The authors acknowledge the families and patients for their participation and thank the numerous health care providers and community clinics for their support and participation in GALA II (4).

The GGAF study is supported by funding from the 5 sources that form GGAF. The AF RISK study is supported by the Netherlands Heart Foundation (grant NHS2010B233), and the Center for Translational Molecular Medicine. Both the Young-AF and Biomarker-AF studies are supported by funding from the University Medical Center Groningen. The GIPS-III trial was supported by grant 95103007 from ZonMw, the Netherlands Organization for Health Research and Development. The PREVEND study is supported by the Dutch Kidney Foundation (grant E0.13) and the Netherlands Heart Foundation (grant NHS2010B280).

We thank contributors who collected samples and/or data used in this study, as well as subjects whose help and participation made this work possible. Biospecimens were collected as part of the eMERGE Network funded by the NHGRI through U01HG8673 to Northwestern University.

This research utilizes resources provided by the Type 1 Diabetes Genetics Consortium, a collaborative clinical study sponsored by the NIDDK, NHGRI, the National Institute of Child Health and Human Development (NICHD), and the Juvenile Diabetes Research Foundation International (JDRF), and supported by U01 DK062418.

Data collection for this project was supported by the McDonnell Genome Institute of Washington University. We thank all participants for their contributions.

We thank contributors who collected samples and/or data used in this study, as well as subjects whose help and participation made this work possible. Biospecimen collection was supported by grants from the Academy of Finland (321428), National Institutes of Health, Sigrid Juselius Foundation, and Finnish Foundation for Cardiovascular Research.

We thank contributors who collected samples and/or data used in this study, as well as subjects whose help and participation made this work possible. Biospecimens were provided by the Emory University School of Medicine.

The dataset(s) used for the analyses described were obtained from Vanderbilt University Medical Center's BioVU, which is supported by institutional funding and by the Vanderbilt CTSA grant ULTR000445 from NCATS/NIH.

We thank contributors who collected samples and/or data used in this study, as well as subjects whose help and participation made this work possible. Biospecimen collection was funded by BG Medicine.

The GeneBank study has been supported by grants from the National Institutes of Health and the Office of Dietary Supplements, including R01HL103866, P01HL147823, and R01HL126827.

This work has been supported by scientific research grants from the Ministry of Education, Science and Culture of Japan (No. 16K19394, 18K08064, and 19K08575), and Japanese Circulation Society (project for genome analysis in cardiovascular diseases). We express our special thanks to the contributors who collected samples and/or data used in this study, as well as subjects whose help and participation made this work possible.

The Centers for Common Disease Genomics are funded by the National Human Genome Research Institute and the National Heart, Lung, and Blood Institute. This work was supported by UM1HG008901.

The ACE2 cohort was collected and phenotyped as part of the Autism Center of Excellence grant MH100027.

We thank contributors who collected samples and/or data used in this study, as well as subjects whose help and participation made this work possible. Biospecimens were provided by the WHI program, which is funded by the National Heart, Lung, and Blood Institute, National Institutes of

Health, U.S. Department of Health and Human Services through contracts 75N92021D00001, 75N92021D00002, 75N92021D00003, 75N92021D00004, 75N92021D00005.

At Vanderbilt, the research was supported by grants from the American Heart Association (11CRP742009, EIA 0940116N) and the NIH (R01 HL092217, U19 HL65962, and UL1 RR024975). The project was also supported by a CTSA award (UL1 TR00045) from the National Center for Advancing Translational Sciences.

Funding was from American Heart Association grant 15SFRN23910002 and the National Institutes of Health grant HL149620.

We thank contributors who collected samples and/or data used in this study, as well as subjects whose help and participation made this work possible. Biospecimens were obtained from the Sao Paulo Brazil Cohort on Coronary Artery Disease, which was supported by institutional funding from the Fundacao Zerbini and by Fapesp grant 2013/12877-4. IRB approval from Cappesq, number 09295712.3.0000.0068.

We thank contributors who collected samples and/or data used in this study, as well as subjects whose help and participation made this work possible. Biospecimens were provided by the SCCS, which was supported by grants from the National Cancer Institute (R01 CA092447) and the American Recovery and Reinvestment Act (3R01 CA092447-08S1).

This project was supported, in part, with support from the Indiana Clinical and Translational Sciences Institute, funded, in part by Grant Number UL1TR001108 from the National Institutes of Health, National Center for Advancing Translational Sciences, Clinical and Translational Sciences Award and the National Center for Research Resources, Construction grant number RR020128 and the Lilly Endowment. We thank contributors who collected samples and/or data used in this study, as well as subjects whose help and participation made this work possible.

The Multiethnic Cohort (MEC) is supported by NIH/NCI grant U01CA164973.
